# Design and clinical validation of a 3D-printed nasopharyngeal swab for COVID-19 testing

**DOI:** 10.1101/2020.06.18.20134791

**Authors:** Joshua K Tay, Gail B Cross, Chun Kiat Lee, Benedict Yan, Jerold Loh, Zhen Yu Lim, Nicholas Ngiam, Jeremy Chee, Soo Wah Gan, Anmol Saraf, Wai Tung Eason Chow, Han Lee Goh, Chor Hiang Siow, Derrick WQ Lian, Woei Shyang Loh, Kwok Seng Loh, Vincent TK Chow, De Yun Wang, Jerry YH Fuh, Ching-Chiuan Yen, John EL Wong, David M Allen

**Affiliations:** Department of Otolaryngology – Head and Neck Surgery, National University of Singapore; Division of Infectious Diseases, Department of Medicine, National University Hospital, Singapore; Department of Medicine, National University of Singapore, Singapore; Molecular Diagnostic Centre, Department of Laboratory Medicine, National University Hospital, Singapore; Centre for Additive Manufacturing, National University of Singapore; Keio-NUS CUTE Center, National University of Singapore, Singapore; Department of Pathology, National University of Singapore, Singapore; Department of Microbiology and Immunology, National University of Singapore, Singapore; Department of Mechanical Engineering, National University of Singapore, Singapore; Division of Industrial Design, National University of Singapore, Singapore; Department of Hematology-Oncology, National University Cancer Institute, Singapore

## Abstract

We describe the development and validation of a novel 3D-printed nasopharyngeal swab for the identification of SARS-CoV-2. We subjected the novel swab to mechanical and fluid absorption testing ex-vivo, and confirmed its ability to retain and release murine coronavirus and SARS-CoV-2. Compared to the Copan FLOQSwab, the novel swab displayed excellent correlation of RT-PCR cycle threshold values on paired clinical testing in COVID-19 patients, at r = 0.918 and 0.943 for the SARS-CoV-2 ORF1/a and sarbecovirus E-gene respectively. Overall positive and negative percent agreement was 90.6% and 100% respectively on a dual-assay RT-PCR platform, with discordant samples observed only at high cycle thresholds. When carefully designed and tested, 3D-printed swabs are a viable alternative to traditional swabs and will help mitigate strained resources in the escalating COVID-19 pandemic.

## Introduction

Since December 2019, the COVID-19 pandemic has spread to 188 countries, with 7.5 million cases worldwide and more than 400,000 fatalities.^1^ One of the World Health Organization’s strategic objectives is to control sporadic clusters and prevent community transmission by rapidly identifying all cases.^2^ The identification of cases is typically performed using material collected from a nasopharyngeal swab, which is tested for SARS-CoV-2 using reverse transcriptase polymerase chain reaction (RT-PCR).

With the surge in cases, testing for COVID-19 worldwide has been limited by shortages of critical components, including RNA extraction kits, PCR reagents, transport media and nasopharyngeal swabs.^3-5^ Here, we share our experience in designing, testing, and clinically validating the utility of a non-flocked 3D printed nasopharyngeal swab for the identification of SARS-CoV-2.

## Results

We first considered the clinical requirements of the swab, in relation to its mechanical, material and biological properties. In particular, while some friction is required to enhance the collection of material from the nasopharynx, sharp edges had to be avoided to avoid trauma and epistaxis. We thus designed the absorbent portion of the swab as a reinforced helix comprising outer blades and inward reverse blades to channel collected material into an internal reservoir ensuring sufficient cellular and extracellular matrix could be collected from the nasopharynx (**Figure 1a,b**). We chose medical-grade resin polymer as the material and allowed flexibility up to 180 degrees to permit maneuverability within the narrow nasopharyngeal space. Post-sterilization, the mechanical properties of the Python swab was tested and benchmarked against the FLOQSwab (Flexible Minitip Flocked Swab, COPAN Diagnostics Inc.). The Python swab had increased tensile strength and flexural strength compared to the FLOQSwab, and was able to tolerate torsional twisting of more than 800 degrees on itself (**Figure 1c**). While its capacity to absorb fluids was inferior to the FLOQSwab, the Python swab was able to release a similar amount of fluid on testing with a roll plate approach (**Figure 1d, Extended Data Figure 1**).^6^

**Figure 1.**
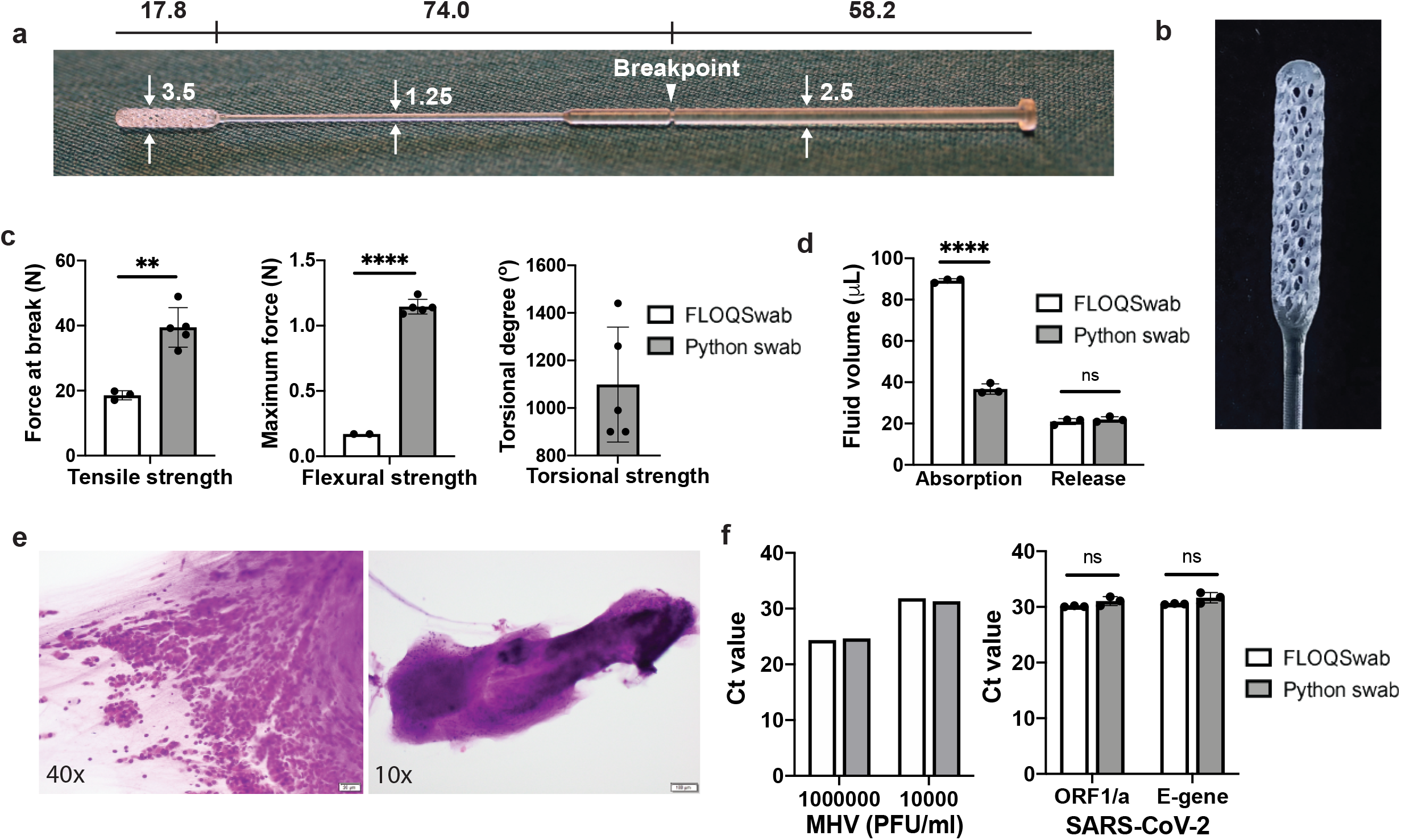
Design and ex-vivo testing of the Python swab. **a**, Overview and approximate dimensions (mm). **b**, High resolution photo of the swab tip. **c**, Mechanical testing of the swab demonstrating its tensile, flexural and torsional strength. **d**, Viscous fluid absorption and release by roll plate approach. **e**, H&E stain of material collected from the swab smeared onto a glass slide, showing sheets of epithelial cells (left) and mucous (right). **f**, RT-PCR results of swabs dipped in murine coronavirus (mouse hepatitis virus, MHV) and SARS-CoV-2 positive viral transport media. ** p < 0.01, **** p < 0.001, ns - not significant, error bars represent one standard deviation.

As with other cytopathic viruses, SARS-CoV-2 invades and multiplies intracellularly, hence the ability of a swab to pick up cellular material is imperative. H&E smears obtained from Python swab samples demonstrate that the swab was able to collect sheets of epithelial cells (**Figure 1e**). We confirmed the ability of the Python swab to retain and release viral samples by dipping swabs in spiked samples with standard inocula of murine coronavirus (mouse hepatitis virus, MHV) and SARS-CoV-2, resuspending in fresh viral transport medium, followed by viral RNA extraction and RT-PCR (**Figure 1f**).

We proceeded then to determine the clinical efficacy of the Python swab in comparison to the FLOQSwab, the standard swab used in our institution, via a case-control study of 40 patients diagnosed with COVID-19 by a positive SARS-CoV-2 RT-PCR test, and 10 control patients with acute respiratory illness who had tested negative for SARS-CoV-2. COVID-19 patients were swabbed at a median of day 8 of illness (range 2 – 14 days). Participants had paired nasopharyngeal swabs through the same nostril performed by trained Infectious Disease and Otolaryngology doctors. The sequence of swabs was randomized based on the sample number assigned to the participant, and the swab technique was standardized. All samples were tested at our institution’s clinical laboratory using a RT-PCR dual assay for the SARS-CoV-2 ORF1/a and the sarbecovirus E-gene. Consistent with our laboratory protocols for clinical samples, samples positive for either of the gene targets were considered to have a confirmed diagnosis of COVID-19.

With the FLOQSwab as the reference, we observed an overall positive percent agreement (PPA) and negative percent agreement (NPA) of the Python swab at 90.6% and 100%, with an overall agreement (OA) of 94% (**Figure 2a**). None of the 10 control samples tested positive. Among 14 COVID-19 cases with swabs performed in the first week of illness, the overall agreement was 100%, while there were 3 false-negative cases out of 26 cases in the second week of illness (**Figure 2b,c**). These 3 false-negative cases tested positive only for the E-gene (and not ORF1/a) on the FLOQSwab, and only at CT values > 37 (**Figure 2d,e**).

**Figure 2.**
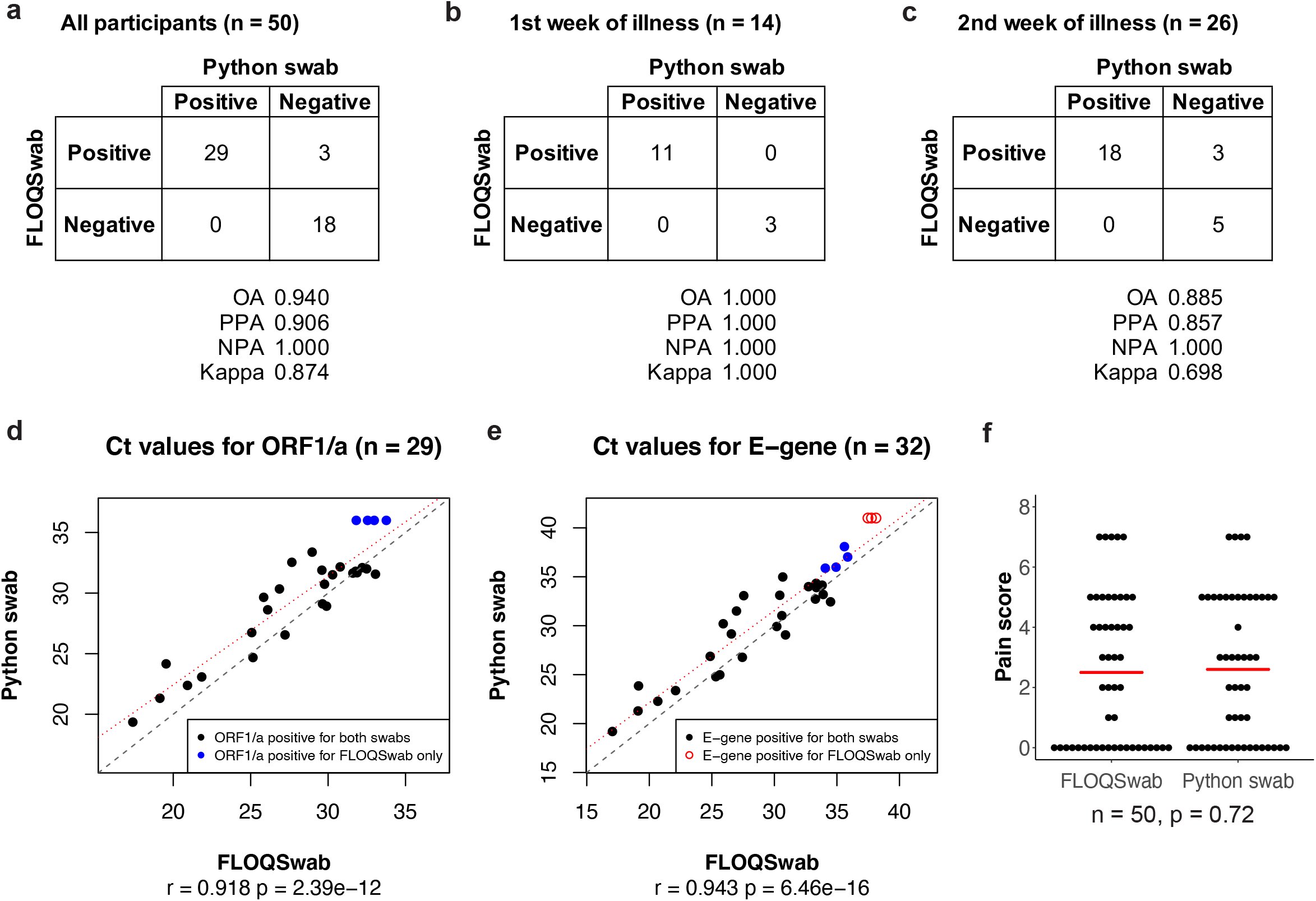
Clinical comparison of Python swab and FLOQSwab. **a-c**, Categorical results for (a) all participants, (b) COVID-19 cases in the first week of illness, and (c) COVID-19 cases in the second week of illness. **d-e**, Correlation plots for all COVID-19 cases that tested positive for ORF1/a (d) and E-gene (e) on RT-PCR. Red line represents line of best fit (linear model). Blue dots represent positive cases that were not identified by the Python swab on ORF1/a, but were identified by the Python swab on E-gene. **f**, Pain score for both swabs, red bar represents the mean for each swab.

There were 4 discordant cases for the Python swab on the SARS-CoV-2 ORF1/a test alone, all of which were positive on the sarbecovirus E-gene for both swabs (**Extended Data Figure 2**). As such, the overall agreement was not affected by these cases in a dual-target testing strategy.

We observed an excellent correlation of Ct values for both swabs, at r = 0.918 (p = 2.39e-12) and 0.943 (p = 6.46e-16) for the ORF1/a and E-gene respectively (**Figure 2d, e**). We also observed that Ct values correlated positively with the day of illness, consistent with a reduction in viral load as a patient recovers (**Extended Data Figure 3**). Mean Ct values for the Python swab were 1.33 and 1.40 higher than the FLOQSwab for the ORF1/a (28.72 vs 27.39, p = 0.0015) and E-gene (30.24 vs 28.84, p = 0.0037) respectively (**Extended Data Figure 4**).

No participant experienced frank epistaxis or swab breakage in this study. There was one episode of blood-stained mucous with the FLOQSwab, and one episode of mild giddiness after the Python swab. There was no difference in mean pain score between the Python swab and FLOQSwab (2.6 vs 2.5, p = 0.72, **Figure 2f**). In terms of comfort, 23 participants (46%) felt no difference between both swabs, 16 (32%) felt that the FLOQSwab more comfortable, while 11 (22%) felt that the Python swab was more comfortable. Overall, 40 participants (80%) found both swabs acceptable, 6 (12%) preferred not to have the FLOQSwab, while 4 (8%) preferred not to have the Python swab.

## Discussion

The accuracy of COVID-19 identification using nasopharyngeal swabs is dependent on a variety of factors, including patient’s viral burden in the nasopharynx, swabbing technique as well as laboratory assays used for testing. This is reflected in a recent meta-analysis showed a pooled sensitivity of RT-PCR testing at 89%, but with a wide range of sensitivities from 50 – 100% between studies.^7^

Although the Python swab had marginally higher Ct values, suggesting that it picked up less material than the FLOQSwab, the overall agreement between both swabs was excellent. It is likely that such small differences in Ct values did not have significant impact on the accuracy of RT-PCR testing, which is highly sensitive for small amounts of material. A strength of our study design is that it allowed us to robustly evaluate the swab by sampling cases with a wide spectrum of viral loads, including those at the limits of detection. Other 3D printed polymer swab designs have been evaluated on COVID-19 cases at diagnosis with false-negative cases observed even at low Ct values of < 25.^8^

In contrast, the false-negative cases in our study had high Ct values (37.48 – 38.12) and were identified only in the second week of disease, these may represent recovering cases with low or no infectivity. Indeed, all three cases were discharged to community care facilities 1 – 4 days after the study swab was performed. A recent study demonstrated that virus could not be isolated in cell culture from patients who had E-gene Ct values ≥ 34, suggesting a lack of infectivity.^9^ Another study observed cell culture infectivity only at Ct values < 24 and < 8 days of illness.^10^ Similarly, in a multicenter cohort of 73 COVID-19 patients in Singapore, no viable virus could be isolated in cell culture when the Ct value was 30 or higher.^11^

Here we show that with careful design and testing, non-flocked 3D-printed polymer swabs can be highly accurate. The Python swab is safe and acceptable for patient use. It is a viable alternative to traditional swabs and will help mitigate strained resources in the escalating COVID-19 pandemic.

## Methods

### Swab design

Nasopharyngeal (NP) swabs should capture respiratory epithelial cells and mucus (cellular-mucus matrix), retain the cellular-mucus matrix while relaying the swab to a transport container, then release the cellular-mucus matrix into the transport media from which viral RNA can be detected in order to be effective. Intuitively, the greater the surface area of the NP swab, the greater the cellular-mucus matrix volume captured and released, with resultant increased sensitivity of the assay. With that goal in mind and with awareness of the diameter of the nasal passage through which the NP swab must journey, a square shaped, helix design with a reservoir concept was selected. The helix makes 1.5 turns from start to end. This allows for surface manipulation (sculpting, flexing and weaving) while retaining sufficient structural integrity for withstanding pushing and turning impact forces. The gaps between the helix blades and the offset of the blades’ inner aspects allow the swab to collect the cellular-mucus matrix via capillary effect and deposit it into the central reservoir. The cellular-mucus matrix contained within the reservoir and adherent to the swab tip is released when the NP swab tip is submerged and agitated in viral transport media.

### 3D-printing and sterilization

The 3D printed swabs were designed and developed using Rhino 6 for Windows (Rhinoceros®, Robert McNeel and Associates, Seattle, WA). After design development, the CAD files were exported into a STereoLithography (.stl) file and printed by the Formlabs Form3 printer (Formlabs Inc., Somerville, MA, USA), and fabricated in Surgical Guide Resin (Formlabs Inc., Somerville, MA, USA) which is non-cytotoxic, not a sensitizer, non-irritating, and complies with international standards (ISO 109933-1:2018 Biological evaluation of medical devices). The 3D printed swabs were then steam sterilized according to recommendations from the United States Centers for Disease Control and Prevention (30 minutes at 121 °C / 250 °F in a gravity displacement autoclave) prior to the testing.

### Tensile, flexural and torsional strength testing

Mechanical testing was performed by an independent testing facility (TÜV SÜD PSB Singapore). 3D printed swabs samples were tested for tensile and flexural strength in accordance with international standards (ISO 527-1:2012 Plastics – Determination of Tensile Properties and ISO 178:2010 Plastics – Determination of Flexural Properties). For tensile testing, swabs were gripped at 10mm from end of the tip. Torsional testing was performed by gripping the swab at 12.7mm from both ends and at a torsional speed of 8 rpm in a clockwise twisting direction. The resulting number of turns (in degrees) when the specimen breaks was reported.

### Fluid absorption and release

Two types of medium were used for the testing, including the Universal Viral Transport Medium (Becton, Dickinson and Company, MD, USA #220527) and a viscous fluid that mimics the human mucus^12^ – 25% (w/v) Pluronic F127 aqueous solution (Sigma Aldrich #P2443), the viscosity of which ranges from 0.6-24 Pa at 20°C from an internal pre-test. Fluid (3ml) was transferred to a 15ml centrifuge tube, which was then weighted using an analytical balances (Sartorius Entris, Göttingen, Germany). The swab was immersed in the fluid for 5 – 10 seconds. The tube was weighted again after removing the swab for the calculation of the absorbed liquid, which is the difference between the weight of the tube before swab immersion and after removing the immersed swab.

Two release methods were performed after fluid absorption, including vortex approach and roll plate approach. The vortex approach is a commonly used procedure to mix an experimental sample and diluent.^13^ An empty 15ml centrifuge tube was weighted; after the immersed swab was placed into the tube, it was vortexed on a lab vortex mixer (Vortex Genie 2, model #G-560, Scientific Industries, USA) for 5 sec. The swab was then removed and the tube was weighted again; the difference of the tube weights represents the release liquid. The roll plate approach provides a semi moisture environment that is frequently used in laboratory and clinical testing for swabs.^6^ A swab was weighted before the absorption. After the absorption procedure, the swab was transferred and rolled with exerting downward pressure on a 1.5% agar plate (Sigma Aldrich #05040); the swab tip was rolled back and forth across the agar surface. Then, the swab was weighted again. The difference of the weight of the swab before and after the roll plate procedure was used as the release weight of liquid. The volumes of the absorption and release were calculated using the weights and the density of the two liquids, which were 0.00103 g/μl (universal viral transport media) and 0.00102 g/μl (viscous fluid) from an internal pre-test. The experiment was repeated three times for each composition of the swab types, liquids, and release methods.

### Murine coronavirus testing

MHV (strain A59) concentrations of 10^6^ and 10^4^ plaque-forming units (PFU) were each freshly prepared in a microfuge tube containing 1 ml of Dulbecco’s modified Eagle’s medium (DMEM). Into each spiked sample was dipped each swab, with swirling and twisting of the swab head for 10 sec, before transferring the infected swab to a fresh tube containing 1 ml DMEM. From the latter, 140 μl was obtained for viral RNA extraction using the QIAamp Viral RNA Mini kit (Qiagen, Germany). Each viral RNA sample (300 ng) was reverse-transcribed to cDNA in a volume of 12.5 μl comprising M-MLV reverse transcriptase (Promega, USA), and MHV-specific primers MHV-NF (5’-ACGCTTACATTATCWACTTC-3’) and MHV-NR (5’-GATCTAAATTAGAATTGGTC-3’). Each cDNA sample (1 μl) was subjected to real-time PCR using FastStart Essential DNA Green Master reaction mix (Roche, Singapore) together with MHV-NF and MHV-NR primers targeting a 256-bp fragment of the MHV N gene. Negative controls without cDNA template were also included. Thermal cycling was conducted using the LightCycler 96 Real-Time PCR System (Roche, Singapore), with the following parameters: pre-incubation at 95oC for 5 min, followed by 55 cycles each at 95°C for 10 sec, 40°C for 5 sec, and 72°C for 8 sec. The relative efficiency of the test swabs was compared based on the determined threshold cycle (Ct) values.

### Human samples

Between May 11 and May 12, 2020, we prospectively recruited 40 adults with a laboratory-confirmed diagnosis of COVID-19 admitted to a tertiary healthcare institution in Singapore. Only adults aged between 21 – 90 years with a nasopharyngeal swab positive for SARS-CoV-2 on RT-PCR were eligible. Patients with known bleeding diathesis were excluded. 10 adults admitted for acute respiratory illness tested to be negative for SARS-CoV-2 were recruited as controls.

All participants were recruited with informed consent, and the study was approved by the Institutional Review Board of the National Healthcare Group, Singapore (Approval number: DSRB 2020/00464). All parts of the study were performed in compliance with the Human Biomedical Research Act, Singapore.

Demographic information such as age, gender and race, as well as day of illness was recorded. Participants were subjected to two consecutive nasopharyngeal swabs in the same nostril using the COPAN FLOQSwab (Becton, Dickinson and Company, MD, USA #220527) and the novel Python swab. Participants assigned an even case number received the FLOQSwab first, followed by the Python swab, while the reverse was true for participants assigned with an odd case number. Swabs were performed in full PPE (personal protective equipment) by trained Infectious Disease or Otolaryngology doctors who routinely administer nasopharyngeal swab testing in our institution during the COVID-19 pandemic. For each participant, both swabs were administered by the same doctor. The number of turns for each swab was standardized at five to ten 360° rotations, with consistency in number of rotations between both swabs emphasized during the study initiation briefing. Immediately after each swab was taken, it was placed into 3ml of BD Universal Viral Transport media (Becton, Dickinson and Company, MD, USA #220527) typically used for FLOQSwabs.

All COVID-19 cases were swabbed within the first 14 days of onset of symptoms. A copy of the study protocol is available in the supplementary information accompanying this article. While the study protocol allowed for either nasopharyngeal swabs or oropharyngeal swabs to be performed, this was standardized to only nasopharyngeal swabs for consistency.

### Adverse event reporting and acceptability questionnaire

Immediately after the swabs were administered, the pain score, as well as any adverse events including epistaxis and nausea, were specifically recorded for each swab. An acceptability questionnaire was then administered recording the participant’s perception of comfort and discomfort between the swabs, as well as acceptability for future testing.

### Clinical testing for SARS-CoV-2

All swabs were tested for SARS-CoV-2 virus using the cobas SARS-CoV-2 test which is a dual-target qualitative real-time RT-PCR assay for use on the high-throughput automated cobas 6800 System (Roche Molecular Systems, Inc., Branchburg, NJ), according to the manufacturer’s instructions. Briefly, 600 μL of the universal transport media was transferred to a secondary tube for automated sample preparation (nucleic acid extraction and PCR amplification) on the cobas 6800. Positive and negative controls were included in each run. The assay targets the ORF1ab (nonstructural protein) sequence which is specific to SARS-CoV-2 and the E gene (envelope protein) sequence which is highly conserved among sarbecoviruses. Following the manufacturer guidelines for interpretation and our standard protocol for clinical samples, samples which were positive for the E-gene only would be re-tested to confirm the positive result. Samples with a consistently positive result on the E-gene only were considered presumptive positive.

### Statistical analysis

Statistical analysis for the mechanical testing and fluid experiments were performed in GraphPad Prism version 8.4.2 (GraphPad Software, La Jolla California USA). To compare tensile strength, flexural strength and fluid absorption/release volumes between swabs, an unpaired t-test was used. P-values less than or equal to 0.05 were considered as significant.

Statistical calculations for the clinical study were performed in R version 4.0.0 (R Foundation for Statistical Computing, Vienna, Austria). Power calculations were performed using the R package: Basic Functions for Power Analysis (pwr), v1.3-0. To identify a modest correlation between the results of both swabs at r = 0.5, 28 COVID-19 positive participants would be required (power 0.8, significance = 0.05, two-sided). Assuming 75% of COVID-19 positive cases remain positive for SARS-CoV-2 during the first 2 weeks of illness, a minimum of 38 COVID-19 positive cases would have to be recruited.

Positive and negative percent agreement were calculated for each RT-PCR target based on 2 x 2 contingency tables, using the categorical results from the FLOQSwab as the reference standard. Overall accuracy was determined by the fraction of true positives and true negatives. Cohen’s kappa was used to measure the agreement between the categorical outcomes from both swabs.

Pearson’s correlation coefficient was used to evaluate the correlation of RT-PCR cycle threshold values between both swabs. For discordant samples (positive on only one swab), a Ct value of 36 and 41 was assigned for the ORF1/a and E-gene respectively, corresponding to the limit of Ct values observed on the Roche COBAS platform in our institution.

Paired Student’s t-test was used to compare the mean Ct values and pain score between both swabs. P-values less than or equal to 0.05 were considered as significant. Bland-Altman plots were prepared using the paired.plotBA function from the R package: Paired Data Analysis (PairedData), v1.1.1.

## Data Availability

The data shown in the manuscript is available upon request from the corresponding author. The source code to reproduce the clinical statistical analysis is made available at https://github.com/josh-tay/COVID-19_Python_swab.

## Acknowledgements

We acknowledge the support of our medical colleagues and patient participants during the clinical study, as well as the National University Hospital Operating Theatre for their support in sterilizing the Python swabs. We are grateful to Dr. Ho Chaw Sing and the National Additive Manufacturing Innovation Cluster (NAMIC) Singapore, for their coordinating efforts between stakeholders and regulatory agencies. Images of the final swab were also contributed by Eye-2-Eye Communications and Structo. Data from the mechanical testing was kindly provided by TÜV SÜD PSB Singapore.

## Author contributions

Conceptualization of study DMA, JEW, JYF, CCY, DYW. Swab design, mechanical testing and fluid testing CCY, JYF, DYW, DMA, SWG, WTEC, AS. Design of clinical study, recruitment of participants, collection of data JKT, GBC, DMA, WSL, KSL, JL, ZYL, NN, JC, HLG, CHS. Clinical laboratory experiments CKL, BY, VTC, DL. Analysis of the clinical data JKT, CKL, BY, GBC. Drafting of the manuscript JKT, GBC, DMA, CCY, JYF, VTC, SWG, CKL. All authors provided feedback and approved the final draft of the manuscript.

## Extended Data

**Extended Data Figure 1.**
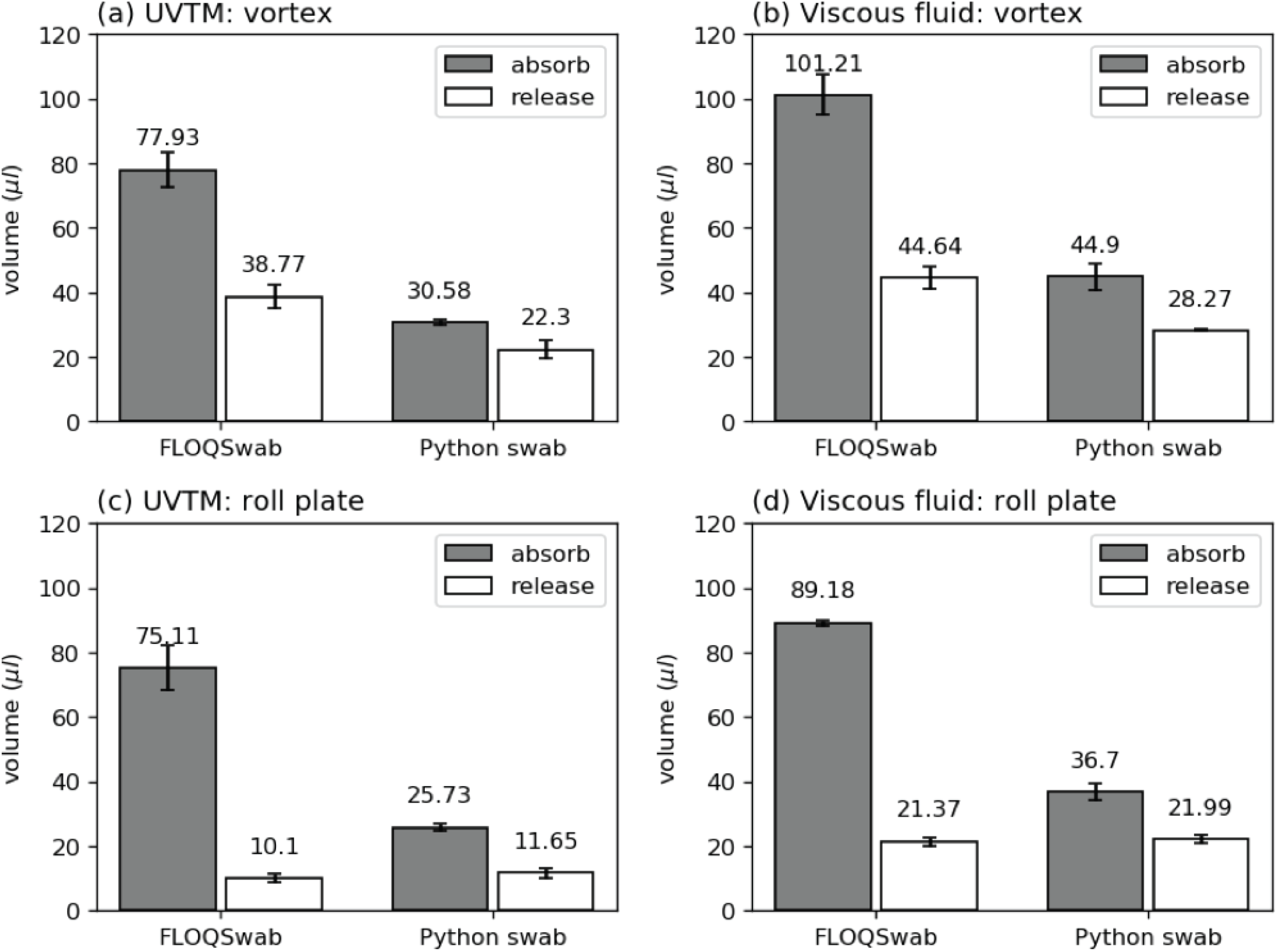
Fluid release and absorption. **a-b**, Fluid absorption and release for the FLO-QSwab and Python swab for (**a**) universal viral transport media (UVTM) and (**b**) viscous fluid (25% w/v Pluronic F127 aqueous solution), tested using the vortex approach. **c-d**, Fluid absorption and release for the FLOQSwab and Python swab for (**c**) UVTM and (**d**) viscous fluid, tested using the roll plate approach.

**Extended Data Figure 2.**
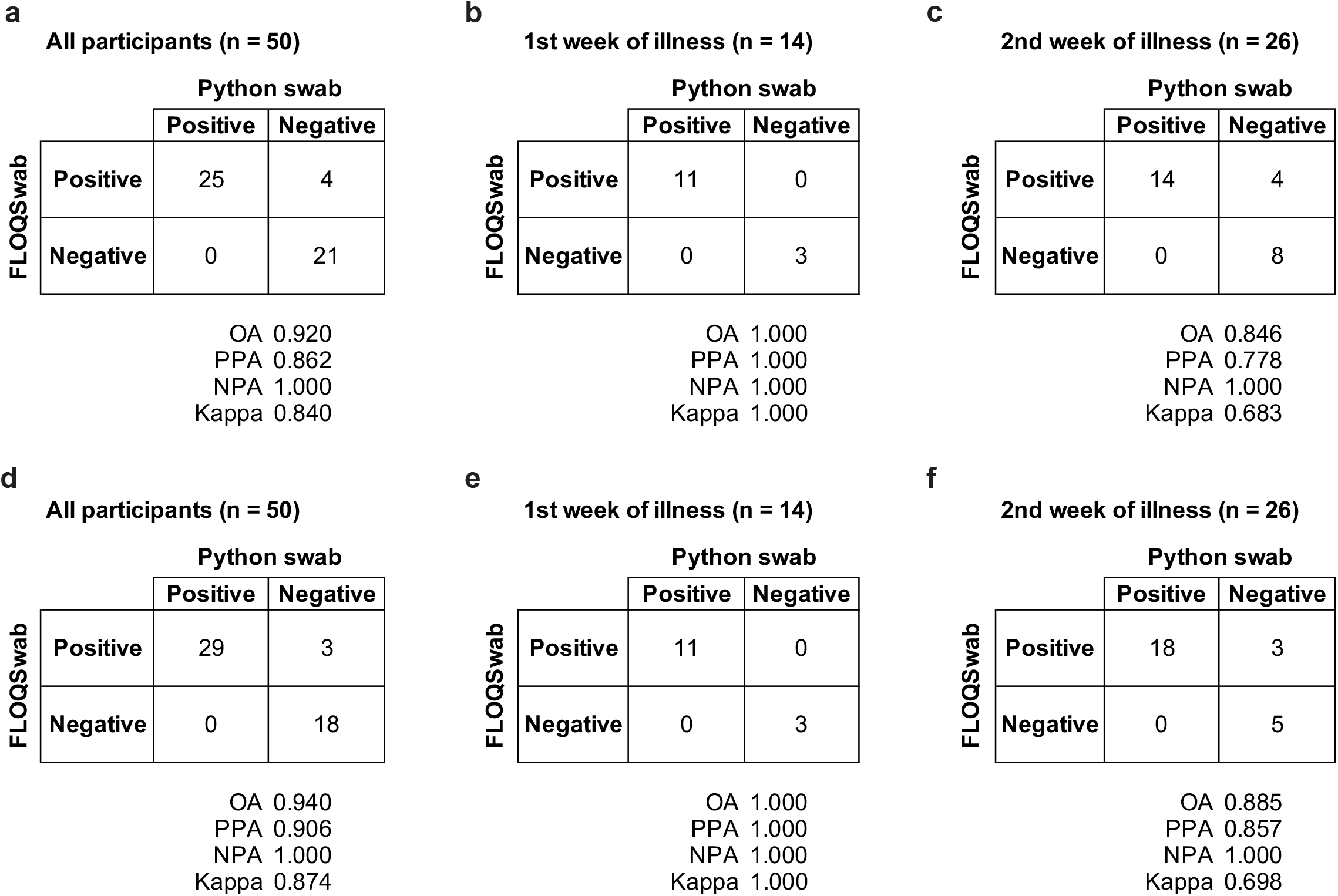
Categorical results comparing the Python swab and FLOQSwab for ORF1/a **(a-c)** and E-gene **(d-f)** targets.

**Extended Data Figure 3.**
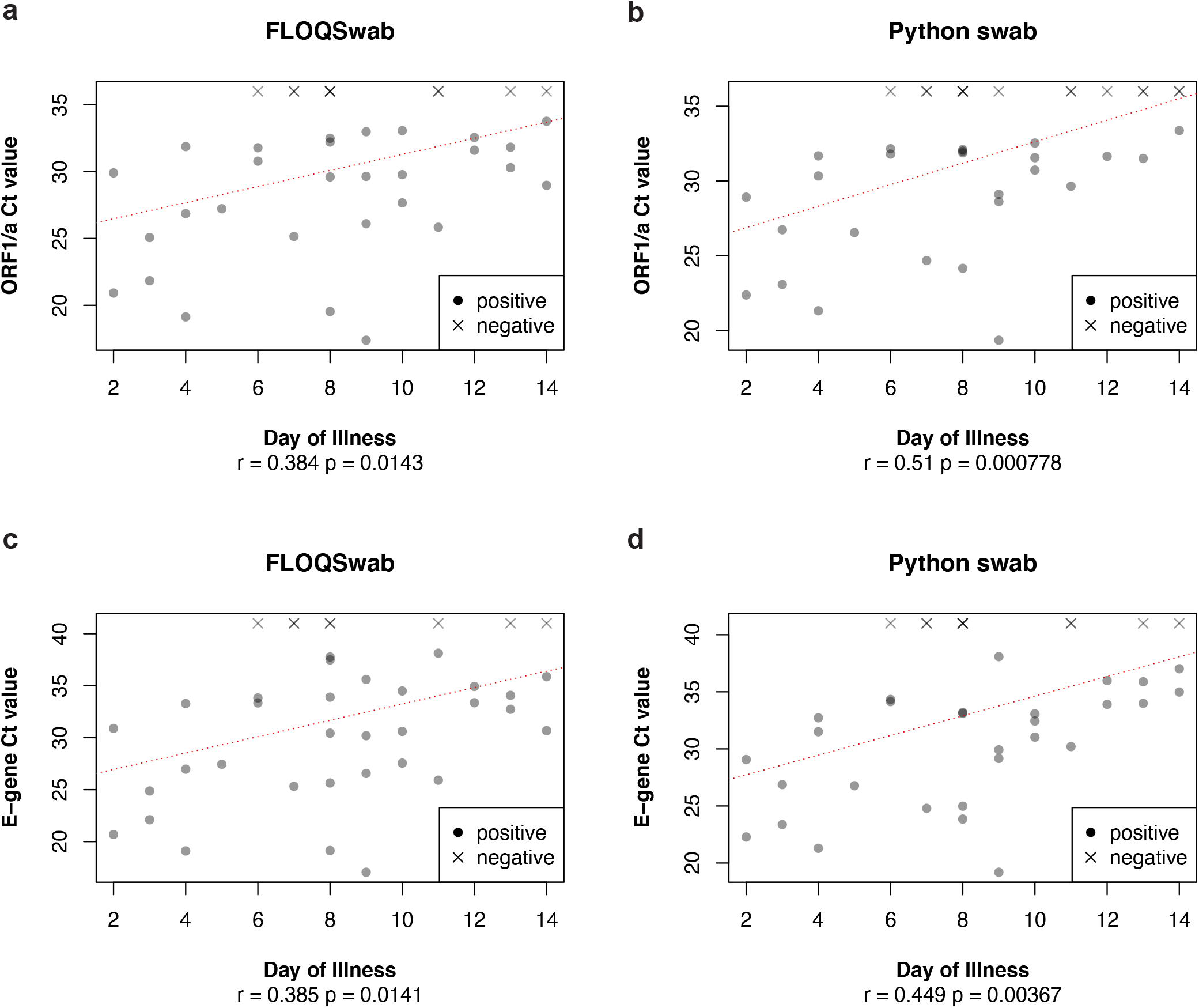
Ct values and day of illness for the ORF1/a **(a,b)** and the E-gene **(c,d)**. Red line represents line of best fit in a linear model.

**Extended Data Figure 4.**
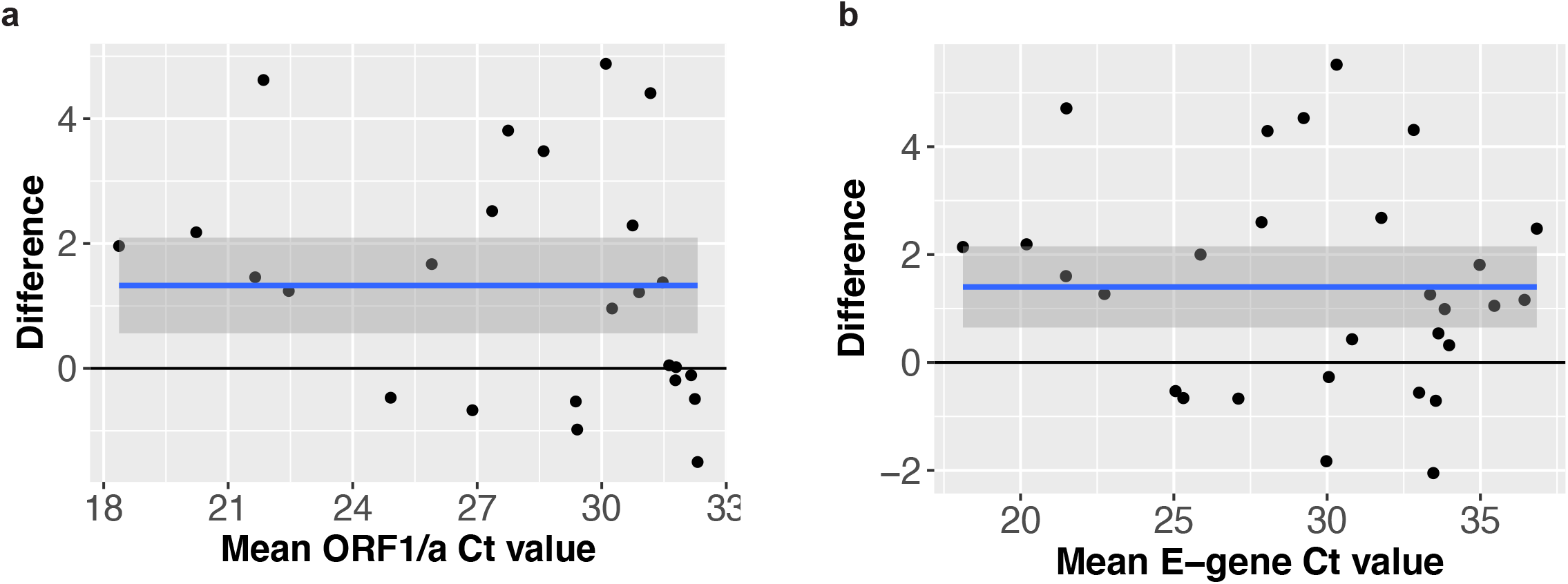
Bland-Altman plots for the comparison of Ct values between paired positive swabs for the ORF1/a (**a**) and E-gene (**b**) respectively. The horizontal axis shows the mean Ct value for each pair of swabs, while the vertical axis shows the difference in Ct value between the swabs (Ct value for Python swab - Ct value for FLOQSwab). The mean difference (+1.33 for ORF1/a and +1.40 for E-gene) is represented by the blue bar and the shaded area represents the boundaries of the 95% confidence interval.

